# Impact of daily octenidine skin washing on antiseptic tolerance of coagulase-negative staphylococci in two neonatal intensive care units with different bathing practices

**DOI:** 10.1101/2022.06.23.22276813

**Authors:** Heather Felgate, Charlotte Quinn, Ben Richardson, Carol Hudson, Dheeraj Sethi, Sam Oddie, Paul Clarke, Mark Webber

## Abstract

**Background:** Coagulase Negative Staphylococci (CoNS) are responsible for 80-90 % of catheter related sepsis in neonates which can cause life threatening and damaging effects. NICUs within the UK use various practices to decolonise neonates to prevent infection ranging from regular full body bathing to localised skin decontamination before insertion of indwelling devices. There is a disparity in bathing practices for infants admitted onto neonatal units, with some choosing to regularly bathe infants and others not, and some routinely washing with skin antiseptics, and others not.

**Aim:** To compare the abundance of CoNS within two UK NICUs with different approaches to skin bathing and to test their tolerances to antiseptics.

**Methods:** A collection of CoNS from two UK based NICUs with differing bathing routines for neonates were collated and tested for susceptibility to the antiseptics in use, octenidine and chlorhexidine.

**Findings:** Regular bathing of neonates in octenidine did not decrease the abundance of organisms on neonatal skin. Isolates from the unit where octenidine was in frequent use did not show any increased antiseptic tolerance. Isolates from the unit where regular bathing was not routine practice were less susceptible to both antiseptics.

**Conclusion:** Frequent whole-body skin washing with octenidine does not appear to result in a lasting reduction in numbers of organisms found on the skin but also does not appear to select for antiseptic tolerant CoNS.

## Introduction

Infection is common amongst premature and low birth weight infants, due to the immaturity of their immune system, skin and mucosal barriers [1, 2]. Late onset infection (LOI), occurring after the first 72 hours after birth, is usually nosocomial and caused by organisms from the skin microbiota or hospital environment [3]. Within neonatal intensive care units (NICUs), invasive procedures are often essential for management but indwelling catheters are a major source of infection [4]. Coagulase-negative staphylococci (CoNS) are common skin commensals, which cause up to 80%– 90% of LOI in NICUs. Catheter-related sepsis can be life-threatening and cause permanent lifelong injury and disability in survivors, including cerebral palsy and other adverse neurodevelopmental problems [3, 5-8].

CoNS rapidly colonise the skin of infants after birth, with the most prevalent species being *S. epidermidis, S. haemolyticus* and *S. warneri* [9]. Disruption of the skin barrier by implantation of intravascular devices can lead to contamination of the outside of these devices (such as central venous catheters [CVC]). This can then lead to bloodstream and catheter-related infections, which can in turn lead to systemic infection and neonatal sepsis [4, 10, 11].

Antiseptics are used pre-implantation to minimise the risk of infection at the site of a skin breach. In addition, within both adult and paediatric populations, there is evidence that regular bathing using antiseptics including chlorhexidine gluconate (CHX) can reduce the number of hospital acquired infections within intensive care [12-14]. However this has not been observed for CHX-based body washing in neonates[15]. Whilst there are national evidence-based guidelines for antiseptic use in children, there is no UK guidance in place for infants who are less than two months old [16]. Due to this lack of UK standardised guidance for topical antiseptic use within NICUs, there are a large range of practices in operation, from regular full body bathing to just local site decontamination before insertion of indwelling devices[17]. There is also a wide disparity between different antiseptics and frequency of bathing, with the most common antiseptics used being octenidine (OCT), povidone iodine and alcohol-/aqueous-based CHX which is frequently used in widely varying concentrations depending on hospital protocols [18, 19].

CHX is a cationic bisguanide with a broad spectrum of antimicrobial activity [20]. It has been shown that regular bathing with CHX significantly reduces the bacterial skin burden in neonates [21], however, the duration of this reduction and subsequent impact on reducing neonatal bloodstream infections and sepsis, is much less clearcut [8, 22, 23]. OCT is a bis-pyridine compound which also has a broad spectrum of antimicrobial activity. Very few studies have examined the use of OCT within a neonatal population, however there is evidence that it is effective at reducing hospital acquired infection amongst adults and older children [24-26]. OCT has been introduced as a body wash as it is reportedly mild and suitable for patients with vulnerable skin.

In this work, we compared antiseptic susceptibility of isolates of CoNS from two UK NICUs; one unit carries out regular whole-body washes for infants, using an OCT based antiseptic (Bradford Royal Infirmary). The other NICU (Norfolk and Norwich University Hospital) does not routinely bathe infants between admission and discharge.

Our hypothesis was that CoNS isolates from the skin in infants who undergo daily whole-body skin washing with OCT will show higher MIC to OCT compared to infants who were not routinely bathed. Thus primary outcome should be the aim to determine whether routine washing of babies with OCT impacted the abundance of CoNS isolated from skin, and impacted tolerance to OCT. Secondary aim was to see if regular washing of babies with OCT impacted tolerance to CHX compared with in CoNS isolates from infants who were not routinely bathed.

### Aims

To determine whether routine washing of babies with OCT may impact the abundance of CoNS isolated from skin, and tolerance to OCT or CHX.

## Materials and Methods

### Study Sites and routine cleansing practices

This work involved NICUs in two hospitals; at the Norfolk and Norwich University Hospital (NNUH) bathing or whole-body skin cleansing was not practised routinely on infants. At the Bradford Royal Infirmary (BRI), infants ≥ 26 weeks were washed daily using Octenisan® (0.3 % octenidine) which was applied to the skin using cotton wool and then washed off water. Infants <26 weeks or with broken/ immature skin were excluded from the regime as per hospital policy. Infants ≥27 weeks corrected gestationl age were eligible for inclusion in this study.

Both centres routinely used locally-applied CHX-based antiseptics (0.015% – 2% CHX in 70% isopropanolol) for pre-procedural skin disinfection before the insertion of indwelling catheters.

### Isolate collection

As part of a previous surveillance study from this laboratory in December 2017 to March 2018[27], a panel of ∼800 CoNS were isolated from skin swabs taken at the NICU of the Norfolk and Norwich University Hospital (NNUH). Swabs were taken on admission and once weekly from each baby throughout their NICU stay from various body sites including the ear, axilla, groin and rectum.

Infants admitted to the BRI NICU also had skin swabs taken on admission and then once weekly for their duration of stay, over a period of 8 weeks (between January and March 2020). All infants admitted to the BRI NICU were eligible for inclusion, regardless of gestational age or expected duration of stay. A single charcoal swab (Amies Charcoal Transport Swab) was used to take a body sweep, incorporating the ear, neck, an axilla, umbilical area and groin. The swabbing was typically carried out 12-16 hours after washing occurred. Swabs were stored locally at 4 °C. Batches were securely packaged and posted to the Quadram Institute Bioscience (QIB), Norwich, every 3 weeks, where they were stored at 4 °C upon arrival.

A unique study ID was allotted to each infant enrolled using their anonymised code generated by the BadgerNet neonatal platform (CleverMed, UK). Birth weights, dates of admission and swabs, birth gestational age, gender of infant, birthing method, location of birth and corrected gestational age at enrolment was collected. No identifying data were transmitted out of the participating sites and completed anonymised data were collated at QIB into a master database.

### Isolation of CoNS

Charcoal swabs were streaked on Columbia Blood Agar (CBA; Oxoid Thermo Fiser Scientific, USA), candidate CoNS were then sub-cultured on Mannitol-Salt Agar (MSA; Oxoid Thermo Fiser Scientific, USA). Isolates were tested for coagulase (Coagulase Test Slides, Millipore, Sigma), and any isolates suspected to be *Enterococci* were grown on Bile Aesculin Agar (Oxoid Thermo Fiser Scientific, USA). Finally, catalase tests were used with 20 % hydrogen peroxide. Isolates considered to be CoNS based on the phenotyping above were saved and given a unique study number.

### Antimicrobial susceptibility testing

The minimum inhibitory concentration (MIC) of OCT and CHX was determined for all isolates according to European Committee on Antimicrobial Susceptibility Testing (EUCAST) guidelines[28]. Muller Hinton (MH) Agar (Oxoid) was prepared with concentrations of antiseptics ranging from 0.25 µg/mL to 64 µg/mL. Overnight cultures grown in MH broth were diluted 1/10,000 and 1 µL drops were plated on to the antiseptic containing MH Agar and incubated at 37 °C for 24 hrs. Two control strains, TW20 and F77, were used throughout[29]. An MIC breakpoint of 4 µg/mL has been suggested to determine CHX resistance; no breakpoints have been proposed for OCT to date although 2 µg/mL has been used previously as an epidemiological cut off [30].

### Statistics

Data were analysed using GraphPad (PRISM 5). Correlation analysis used nonparametric Spearman tests, one-tailed with confidence levels of 95 %. The nonparametric one-tailed T-Test and the Mann-Whitney test were used to identify significant differences between MIC data with a 95 % confidence level.

### Ethics

The study protocol was reviewed by the Research Services Manager of the Norfolk and Norwich University Hospitals NHS Foundation Trust and was approved as a surveillance study that did not require a formal ethics committee review.

## Results

### Isolation of CoNS from Bradford neonates

A total of 56 infants from BRI were enrolled in the study. From these infants, 200 skin swabs were transported to QIB. These were made up of both admission swabs (n=30) and swabs taken weekly (n=170). One swab was discarded due to inadequate labelling. After swabs were incubated plates typically demonstrated heavy growth of both Gram-positive and Gram-negative isolates demonstrating various colony morphologies. This contrasts with the results in our previous study from NNUH where individual swabs generally had a smaller number of isolates with less mixed cultures. Of 180 Gram-positive isolates, a total of 78 were confirmed as CoNS and retained for phenotypic testing.

### Susceptibility of Bradford CoNS isolates to antiseptics

The MICs of CHX and OCT were determined for the isolates from BRI. Isolates were generally very sensitive to OCT and the MICs ranged between ≤0.125-1 µg/mL with the majority (48.7%) of the isolates (n= 38) being inhibited by ≤0.125 µg/mL. Two isolates had a MIC of 0.25 µg/mL, 32 isolates (41%) had a MIC of 0.5 µg/mL and the remaining 6 (7.7%) had a MIC of OCT of 1 µg/mL.

The isolates were also tested against CHX. For 44.9 % of the isolates (n=35) the MIC of CHX was ≤0.125 µg/mL, for 29 (37.2%) it was 0.25 µg/mL, 1 isolate had a CHX MIC of 0.5 µg/mL and the remaining 12 (15.4 %) had a CHX MIC of 1 µg/mL. No isolates from BRI was above the proposed breakpoints for either antiseptic.

The CHX and OCT MIC data for each of the Bradford isolates were compared against each other to determine whether there was any relationship between the susceptibility to the two agents. This analysis (Figure 1) showed no direct relationship between susceptibility to the two antiseptics (P = 0.4), which is similar to our previous findings[29].

**Figure 1:**
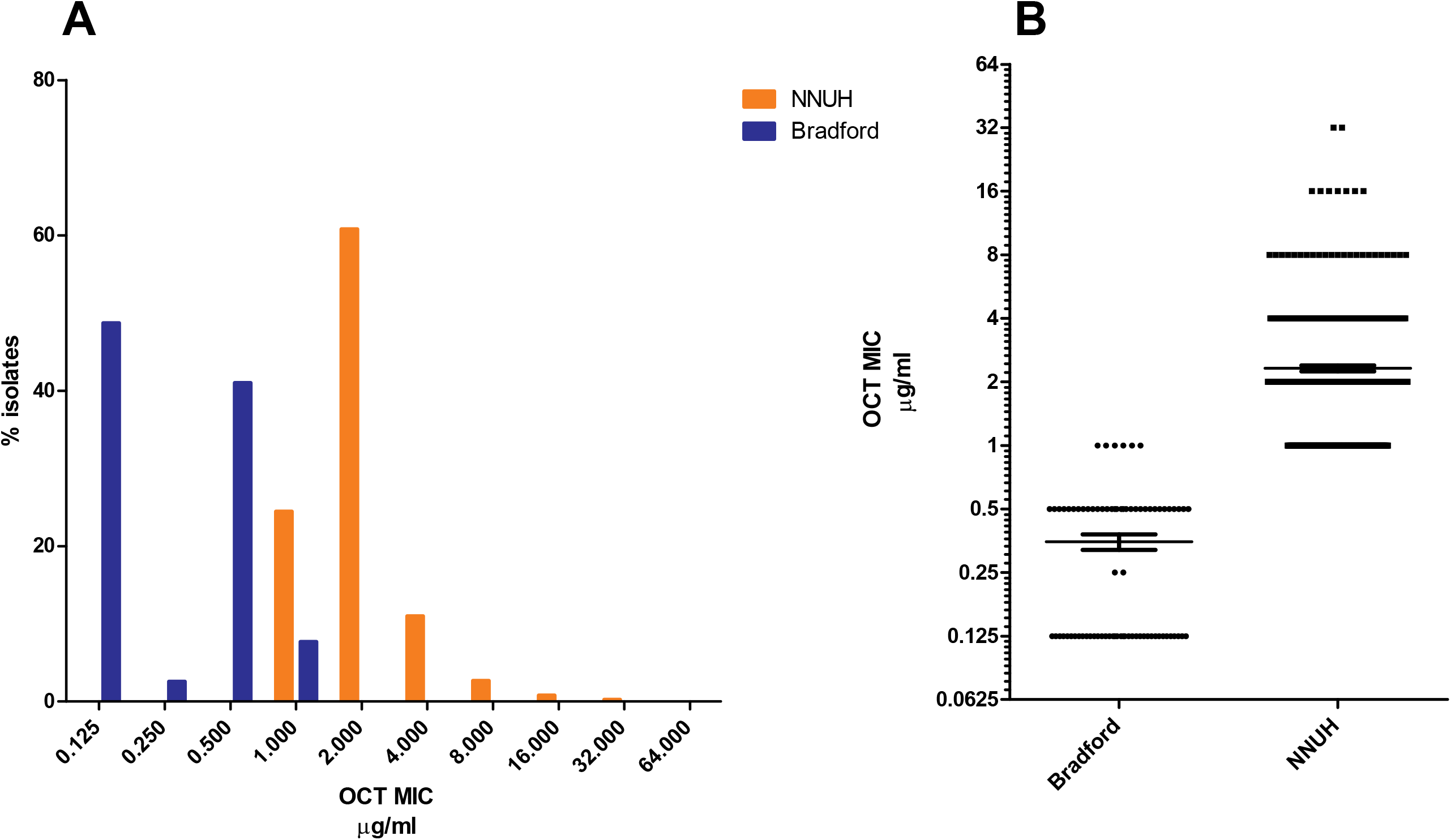
Susceptibility of isolates from BRI to OCT and CHX showed no correlation (p= 0.4, according to Spearman test).

### Comparative antiseptic susceptibility of isolates from Bradford and Norwich

Antiseptic susceptibility of the BRI isolates from the daily OCT-washed babies was compared with the panel of 863 CoNS isolated from babies in the NNUH NICU where infants are not routinely bathed with OCT. A comparison in the susceptibility profiles of the population of CoNS from BRI and NNUH showed significantly decreased susceptibility in the NNUH population to both antiseptics (figures 2 and 3).

**Figure 2:**
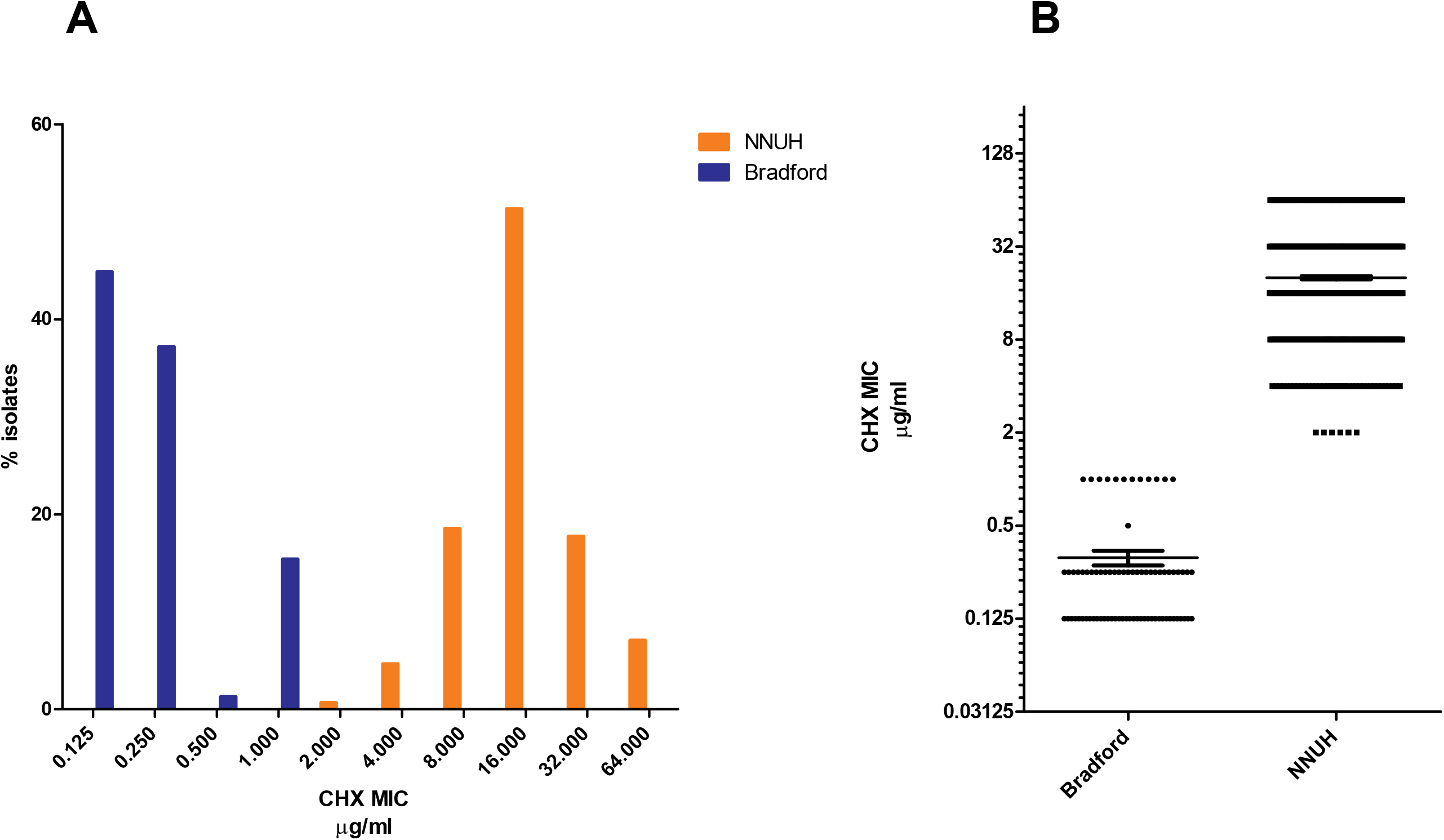
**A** Comparison of MICs of OCT against isolates from BRI where regular daily whole-body OCT washing was in place (n=78) and isolates from NNUH where there was no regular washing of neonates while in NICU (n=863). **B** Boxplot showing numbers of isolates with different OCT MICs from each site (**** *P*= <0.0001). Thin horizontal line indicates the mean and whiskers standard error.

**Figure 3:**
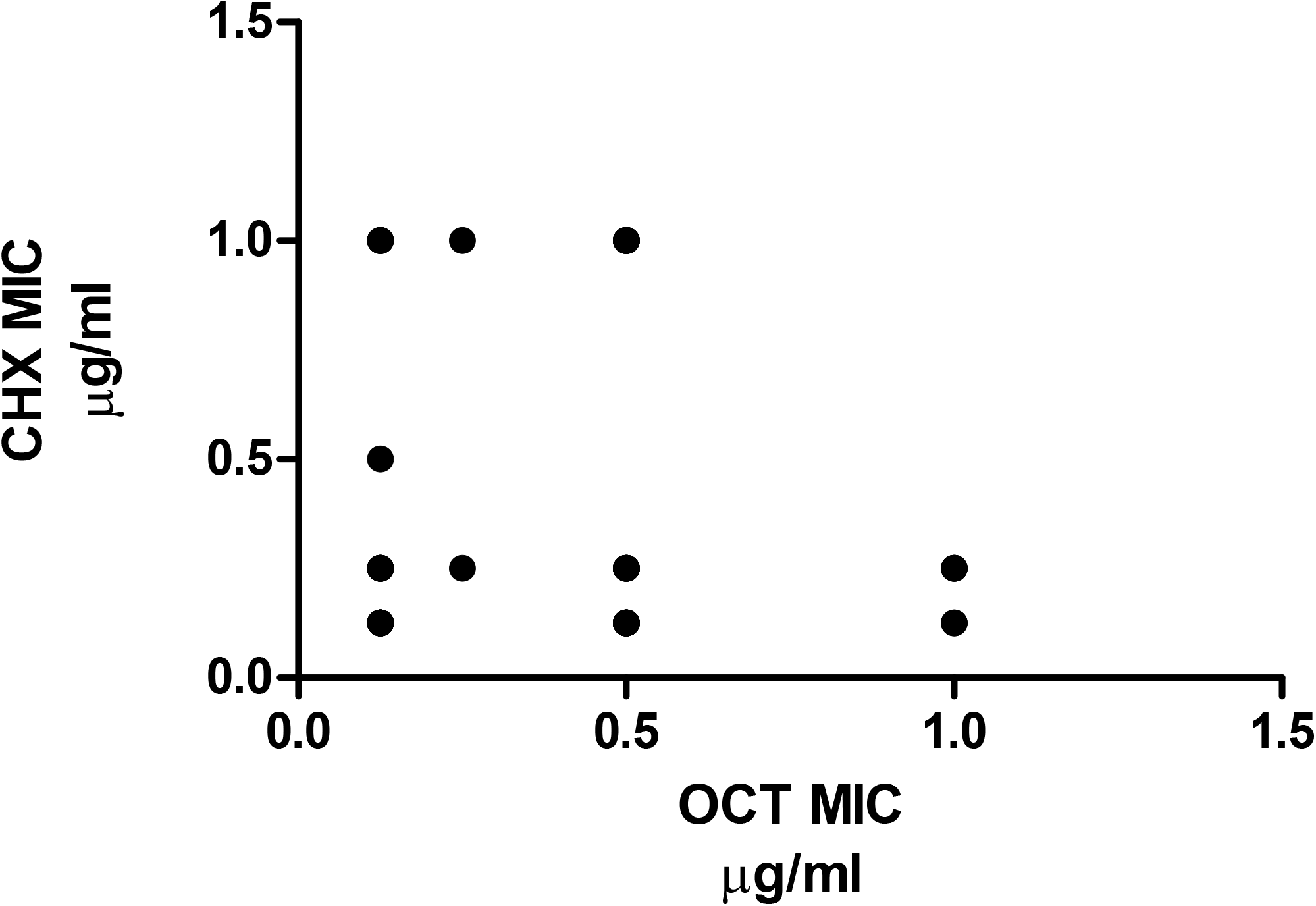
**A** Comparison of MICs of CHX against isolates BRI where regular daily whole-body OCT washing was in place (n=78) and isolates from NNUH where there was no regular washing of neonates while in NICU (n=863). **B** Boxplot showing numbers of isolates with different CHX MICs from each site (**** *P*= <0.0001). Thin horizontal line indicates the mean and whiskers standard error.

The MICs of OCT for infants from NNUH ranged between 1 and 16 µg/mL (mean of 2.319 SEM± 0.078 µg/mL), compared with a narrower range of ≤ 0.125 and 1 µg/mL (mean of 0.394 SEM± 0.029 µg/mL) for BRI isolates (Figure 2a). There was a significant difference in the mean MIC for OCT between Bradford and Norwich NICUs, (*P*=<0.0001, Figure 2B).

The MICs of CHX for NNUH isolates ranged between 2 to 64 µg/mL (mean of 20.1 SEM± 0.5 µg/mL), compared with a range of ≤ 0.125 to 1.0 µg/mL for isolates from babies at BRI (mean of 0.31 SEM± 0.04 µg/mL) (Figure 3). A clear difference in the distribution of CHX susceptibility of the isolates from the two sites can be observed. There was a significant difference between the mean MIC for CHX between the Bradford and NNUH isolates (*P*=<0.0001, Figure 3). In total 817 (94.7 %) isolates from infants at the NNUH had a MIC for CHX greater than 4 µg/mL whereas no isolates from Bradford NICU had a MIC of CHX >1 µg/mL.

## Discussion

In this study we sought to examine the prevalence and antiseptic susceptibility of CoNS from the skin of neonates where washing with OCT is routine daily practice, and to compare the antiseptic susceptibility with a panel of isolates from a unit which does not bathe infants routinely in the period between admission and discharge.

Inoculation of swabs from BRI typically resulted in extensive bacterial growth made up of multiple morphologically distinct bacteria, including a mixture of Gram-positive and Gram-negative bacteria. Therefore, despite being bathed regularly using OCT, the numbers of organisms on the skin of babies from BRI remained high, and in fact more variation was seen than for swabs attained from the NNUH using the same methodology, although this is an anecdotal observation. The skin swabbing technique in BRI involved carrying out a whole-body composite swab, which incorporated the ear, neck, axilla, umbilical area and groin. It has been suggested that there is not much differentiation between differing sites of the body and the skin burden, however this may have contributed towards the high number of organisms recovered, and in particular the greater numbers of putative Gram-negative bacteria [31]. Swabbing was carried out up to 16 hours after the washing of the infants occurred, sampling time was not standardised to fit with practices on the ward, changes over time may have been seen if a defined series of time points were assessed. Regardless of these caveats, it is clear that the OCT washing regime does not sterilise neonatal skin, or that the microbiota is quickly reinstated with multiple organisms soon after washing. Analysis of infants for whom there was both an admission swab and a weekly swab showed a similar number of colonies were picked from both plates - this would argue against acquisition of OCT-tolerant organisms after initial admission.

A previous study showed that for CHX after an initial decrease in the bacterial skin burden after application, the number of recovered organisms increases and baseline levels are reached by approximately 72 hours [21]. CHX demonstrates a ‘substantive effect’, whereby the dried residue of agent remains active in situ for a prolonged period post application onto the skin, which may reflect why a longer period is needed to repopulate the skin microbiota than after OCT exposure.

The susceptibility data did not suggest that repeated/frequent exposure to OCT selects for antiseptic tolerance on skin isolates (Figure 2a and 3a). In fact, isolates from BRI were significantly more susceptible to both antiseptics than those from NNUH and all isolates with highest MICs were from NNUH. This is similar to our recent comparison of the NNUH panel with a German panel (who regularly use OCT based antiseptics for skin decolonization prior to catheter insertion) and again suggests CHX exposure appears more likely to select for antiseptic tolerance than OCT[27]. The substantive effect of CHX may result in long lasting low concentrations of CHX remaining on the skin which might provide an environment for selection of tolerant mutants. Alternatively, CHX is more commonly incorporated in environmental cleaning wipes and products than OCT which may also reflect a greater selective pressure for isolates with decreased tolerance.

### Strengths and Limitations

This is the first study to assess microbiological impacts from practising routine washing of babies with an antiseptic. This study also suggests daily OCT washing does not select for decreased antiseptic susceptibility in CoNS and assessment of the bacterial burden of plates shows OCT washing has a limited impact on any reduction in skin microbiota.

However, limitations in the study mean that possible differences in the genotypes of the strains in circulation between the units were not assessed. The isolation periods between the sites were not exactly contemporaneous although no significant changes in units’ practices occurred in the intervening period. Also a larger number of isolates from NNUH were included which may skew comparisons to some degree.

### Conclusion

In summary, this two-site observational study shows that frequent whole-body skin washing with OCT compared to not routinely washing does not appear to result in a lasting reduction in numbers of organisms found on the skin but also does not appear to select for OCT tolerant organisms. Isolates from the NNUH were much less susceptible to antiseptics than the BRI isolates, suggesting that the bathing of neonates in the BRI NICU does not select for resistance. The data suggest that different antiseptic regimes can have significantly different impacts on the microbiota in terms of both composition and antiseptic susceptibility.

Clinical trials to systematically compare efficacy, safety and microbiological impacts of different antiseptic regimes in order to design evidence informed guidelines are lacking for this vulnerable patient group. Further work on skin cleansing in preventing neonatal sepsis is vital in order to produce best practice guidelines which will minimise infection and potential for selection of antiseptic resistance.

## Data Availability

All data produced in the present study are available upon reasonable request to the authors

## Acknowledgements

We sincerely thank all NNUH and BRI research and clinical nurses who helped with skin swab collection. We are grateful to Julie Dawson, Research Services Manager at NNUH for reviewing our study protocol.

## Conflict of interest

None

## Source of Funding

This work was supported by an award from the Biotechnology and Biological Sciences Research Council (BB/T014644/1).

## Notes

### Competing Interest Statement

The authors have declared no competing interest.

